# Piloting wastewater-based surveillance of norovirus in England

**DOI:** 10.1101/2024.03.11.24303973

**Authors:** David I. Walker, Jessica Witt, Wayne Rostant, Robert Burton, Vicki Davison, Jackie Ditchburn, Nicholas Evens, Reg Godwin, Jane Heywood, James Lowther, Nancy Peters, Jonathan Porter, Paulette Posen, Tyler Wickens, Matthew J. Wade

## Abstract

Wastewater-based epidemiology (WBE) gained widespread use as a tool for supporting clinical disease surveillance during the COVID-19 pandemic. There is now significant interest in the continued development of WBE for other pathogens of clinical significance. In this study, approximately 3,200 samples of wastewater from across England, previously collected for quantification of SARS-CoV-2, were re-analysed for the quantification of norovirus genogroup I (GI) and II (GII). Overall, GI and GII were detected in 93% and 98% of samples respectively, and at least one of the genogroups was detected in 99% of samples. GI was found at significantly lower concentrations than GII, but the proportion of each genogroup varied over time, with GI becoming more prevalent than GII in some areas towards the end of the study period (May 2021 – March 2022). Using relative strength indices (RSI), it was possible to study the trends of each genogroup, and total norovirus over time. Increases in norovirus levels appeared to coincide with the removal of COVID-19 related lockdown restrictions within England. Local Moran’s I analyses indicated several localised outbreaks of both GI and GII across England, notably the possible GI outbreak in the north of England in early 2022. Comparisons of national average norovirus concentrations in wastewater against concomitant norovirus reported case numbers showed a significant linear relationship. This highlights the potential for wastewater-based monitoring of norovirus as a valuable approach to support surveillance of norovirus in communities.

## Introduction

Wastewater-based epidemiological (WBE) has been documented as an approach for monitoring disease since at least the 1930s. Early studies demonstrated that poliovirus could be detected in urban sewage and this could be used to indicate presence of polio in communities (Paul, Trask and Gard, 1940). With the advent of analytical techniques such as PCR, sequencing, and mass spectrometry, quantifying and characterising biological and chemical signatures in wastewater has become simpler, faster, and more precise (Gracia-Lor *et al*., 2017; Sims and Kasprzyk-Hordern, 2020; Kasprzyk-Hordern *et al*., 2023). This has greatly expanded the capacity to investigate and respond to public health threats through the analysis of wastewater samples, which established WBE as a complementary source of data during the COVID-19 pandemic (Diamond *et al*., 2022).

The rapid expansion of the use of WBE tools from the start of the COVID-19 pandemic in early 2020 resulted in wide-scale implementation of wastewater monitoring networks across many countries (COVIDPoops19, 2023; Keshaviah *et al*., 2023). From 2020 to 2022, the Environmental Monitoring for Health Protection (EMHP) programme in England was one of the largest of such networks, with over 500 sites being monitored for SARS-CoV-2 several times per week at its peak (UKHSA, 2023a), covering approximately 74% of the English population (UKHSA, 2023c).

Following WBE for monitoring human health threats such as SARS-CoV-2 and poliovirus (Klapsa *et al*., 2022; Link-Gelles *et al*., 2022), there has been significant interest in its application for monitoring other pathogens, including the spread of Antimicrobial Resistance (AMR) (Aarestrup and Woolhouse, 2020), human respiratory viruses (Boehm *et al*., 2023) and MPox (Wolfe *et al*., 2023).

For WBE to be an effective system for disease surveillance, the target pathogens must enter human wastewater. This is predominantly due to the presence of the pathogens in the faeces and urine of infected individuals, although they may also enter the sewage via other bodily excreta (Jones *et al*., 2020). For respiratory diseases such as COVID-19, the pathogens may be present in faeces due to replication of virus within the gastrointestinal tract or through ingestion of mucus from the upper respiratory tract (Hirose *et al*., 2017). This may result in a relatively low abundance of respiratory pathogens, leading to limited detectability where cases within communities are low, where faecal shedding occurs in a small proportion of infected individuals, or where the target pathogen has limited environmental stability.

In contrast, enteric pathogens such as norovirus (NoV) are often shed in very high numbers in the faeces of infected individuals and tend to be relatively stable in the environment (Bosch, Pintó and Abad, 2006; Gholipour *et al*., 2022). NoV are a major global cause of acute gastroenteritis (AGE). With transmission of NoV through contaminated food and water being an important pathway, and the high viral loads shed by infected individuals (8 – 10 log_10_ gene copies (NoV)/ml versus 2 – 7 log_10_ gene copies (SARS-CoV-2)/ml, (Jones *et al*., 2020)), monitoring of wastewater presents an opportunity for collecting data on temporospatial dynamics of the disease in populations.

NoV specific studies from wastewater samples have demonstrated the value in employing WBE to provide epidemiological information on the disease. Several studies have shown that molecular methods, such as RT-qPCR (Zhou *et al*., 2016) and high-throughput sequencing (Kazama *et al*., 2016; Fumian *et al*., 2019), are able to quantify and characterise the diversity of NoV present in wastewater, with genogroups I (GI), II (GII) and the less common IV (GIV) being those associated with human infection.

In the UK and other developed countries, NoV outbreaks tend to exhibit well-defined seasonality, with peak case numbers occurring in the winter months. However, during the COVID-19 pandemic, there was evidence of decreased NoV prevalence due to the introduction of COVID-19-related non-pharmaceutical interventions (NPIs) in many countries. For example, a study from oyster production areas in Ireland showed that while NoV was detected in 94.3 and 96.6% of oyster samples in the two winters preceding the COVID-19 pandemic, this value decreased to 63.2% in the winter of 2020 to 2021 (Keaveney *et al*., 2022). Wang *et al*. (2023), showed that the transmission of NoV GII was significantly reduced by the introduction of public restrictions on movement and good hygiene practice in Sweden. After these restrictions were lifted in February 2022, peak NoV GII cases were observed at five times the magnitude of those recorded in 2017. This phenomenon was replicated in other countries, such as the UK, where the reduction in NoV reporting was attributed to multiple factors including changes in ascertainment, testing capacity, access to healthcare, and the impact of NPIs (UKHSA, 2023b).

This study contributed to the Pathogen Surveillance in Agriculture, Food and the Environment (PATH-SAFE) Programme, the overall aims of which were to pilot a improved national surveillance systems for the monitoring foodborne disease (FBD) and antimicrobial resistance (AMR) in the UK. We aimed to increase our understanding of how wastewater-based methods can give insights into the prevalence of NoV in communities to aid epidemiological investigations. Using an existing set of samples previously collected for the EMHP programme, we used a ‘one sample, many analyses’ approach to pilot a more cost effective, non-invasive, multi-agency national surveillance system. Using RT-qPCR assays, we quantified NoV genogroup I (GI) and II (GII) as the most prevalent NoV genogroups known to cause disease in humans.

## Methods

### Site and Sample Selection

Samples of screened, untreated sewage were collected as part of the Environmental Monitoring for Health Protection (EMHP) programme, which was to monitor SARS-CoV-2 in wastewater in England as described by Wade et al. (2022) and Morvan *et al*. (2022). A subset of 3,232 samples, collected between 27^th^ May 2021 and 30^th^ March 2022 from 152 sewage treatment works (STW) across England, were selected (Figure 1). The number of STWs in each of the nine regions of England (ONS, 2017) are shown in Table 1. STWs that served the largest populations (based on population data from the Office for National Statistics) were prioritised so that a greater proportion of the population was covered with the minimum number of samples.

**Figure 1:**
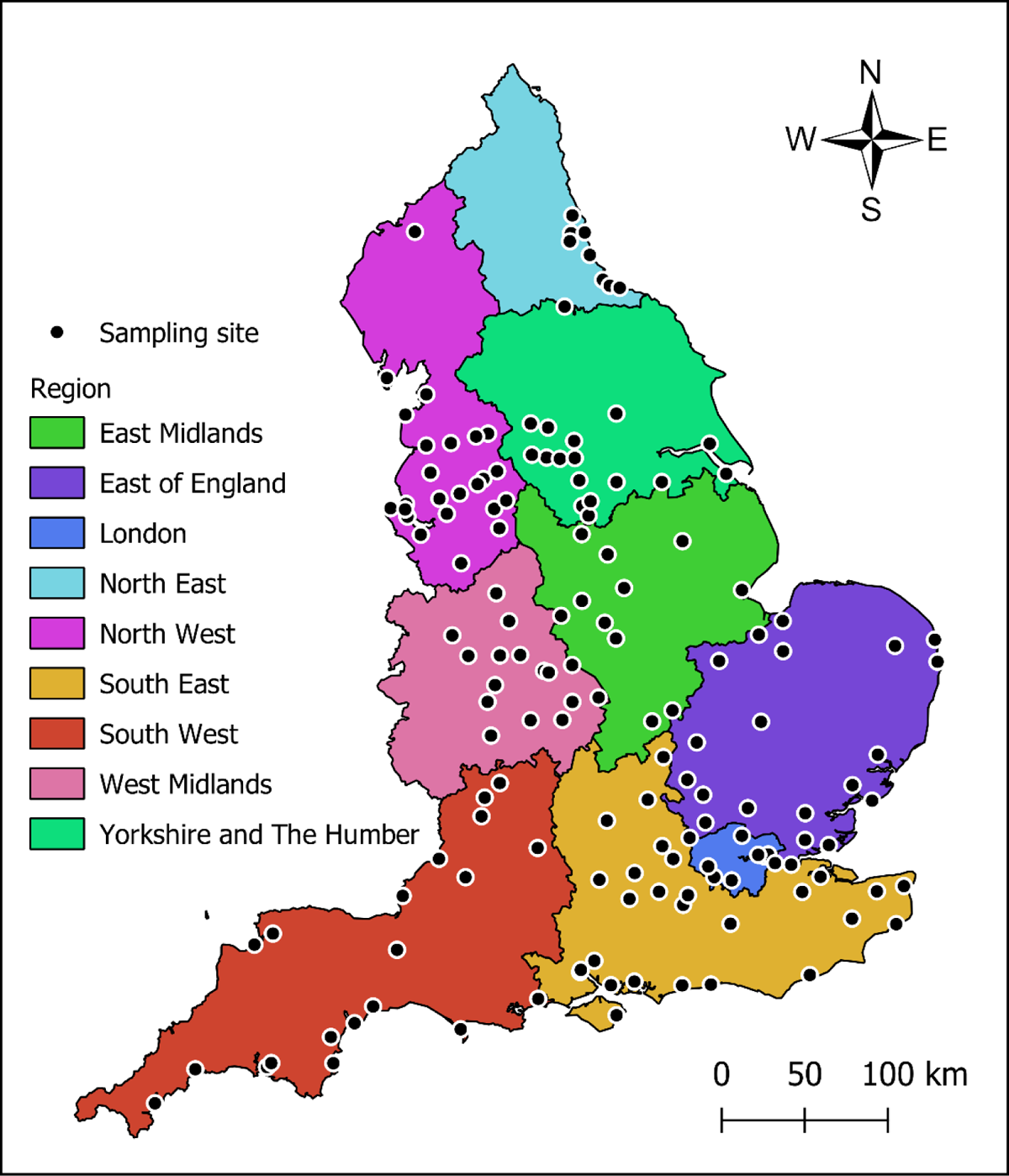
Map showing the sampling sites for this study and the regions of England (ONS, 2017) in which the sites were located.

**Table 1.** Summary of the wastewater samples analysed for norovirus by English region.

| Region | Number of sites | Total samples tested | Earliest sampling date | Latest sampling date | Average sampling interval (days) | Average samples tested per site |
| --- | --- | --- | --- | --- | --- | --- |
| East Midlands | 10 | 221 | 31/05/2021 | 25/03/2022 | 14.2 | 22.1 |
| East of England | 20 | 440 | 31/05/2021 | 21/03/2022 | 13.9 | 22.0 |
| London | 8 | 177 | 30/05/2021 | 25/03/2022 | 13.9 | 22.1 |
| North East | 9 | 172 | 31/05/2021 | 28/03/2022 | 13.9 | 19.1 |
| North West | 24 | 480 | 27/05/2021 | 30/03/2022 | 13.9 | 20.0 |
| South East | 28 | 617 | 28/05/2021 | 30/03/2022 | 13.9 | 22.0 |
| South West | 20 | 429 | 30/05/2021 | 30/03/2022 | 14.0 | 21.5 |
| West Midlands | 17 | 375 | 30/05/2021 | 30/03/2022 | 13.9 | 22.1 |
| Yorkshire and The Humber | 16 | 321 | 30/05/2021 | 25/03/2022 | 14.1 | 20.1 |
| Total | 152 | 3232 | 27/05/2021 | 30/03/2022 | 14.0 | 21.2 |

The STWs selected for this study were estimated to serve a population of approximately 34 million people, equivalent to approximately 60% of the population of England in mid-2021 according to ONS data (ONS, 2022).

We aimed to analyse one sample per STW every two weeks to maximise both the temporal and spatial coverage that could be achieved within the resource limitations of the project. The samples used for this study are summarised in Table 1. Full details of the samples are available from (David I. Walker *et al*., 2024).

### Ammonia and Orthophosphate Quantification

Ammonia and orthophosphate concentrations (mg/l) were measured in wastewater samples within 24 hours of collection according to the Environment Agency’s in-house methods. These methods were based on the relevant Standing Committee of Analysts methods (Standing Committee of Analysts, 1981, 1992) and adapted for use with the Gallery Plus Discrete Analyser (Thermo Fisher Scientific, UK).

### Viral concentration and RNA extraction

Viruses were concentrated and the RNA extracted from wastewater samples according to Walker et al. (2024). Briefly, 200 ml of wastewater was clarified by centrifugation before a phi6 process control and 60 g ammonium sulphate was added to 150 ml of the supernatant and incubated at 4°C for at least 1 hour. These were then further centrifuged, and lysis buffer was added directly to the viral pellet before purification using BioMerieux’s (France) NucliSENS magnetic silica reagents on a KingFisher™ Flex Purification system (Thermofisher).

### RT-qPCR

As part of the COVID-19 monitoring programme, the phi6 process control was used to determine viral recovery as described by Walker et al. (2024). The RNA extracts were subsequently refrozen at −80°C until required for NoV RT-qPCR.

NoV GI and GII were quantified in each sample using a duplex RT-qPCR adapted from Alex-Sanders *et al*. (2023) in duplicate for each sample. The NoV RT-qPCR followed the same procedures as described by Alex-Sanders *et al*., except the GI probe was synthesised by Eurogentec (Belgium) and used an MGB-Eclipse quencher. The GII probe was synthesised by Integrated DNA Technologies (UK) and used a HEX™ reporter dye and was double-quenched by ZEN™ and IB®FQ quenchers. The reactions were carried out on an Aria MX real-time PCR machine (Agilent). Samples were quantified relative to an RNA standard dilution series that was included on each RT-qPCR plate. A duplicate standard curve was constructed from a four point, ten-fold dilution series of RNA Ultramers (Integrated DNA Technology) containing the RT-qPCR target sequences and diluted in Tris-EDTA containing 1% Triton-X (TEX buffer). RT-qPCR results were accepted if the standard curve had an R² of >0.98 and a slope of between −3.6 and −3.1.

The Limit of Detection (LOD) and Limit of Quantification (LOQ) for GI and GII RT-qPCRs were measured based on the methods described by Walker (2022). Previous studies had indicated low RT-qPCR inhibition using this system, so inhibition was not tested in this study (Alex-Sanders *et al*., 2023; Scott *et al*., 2023).

### Data analysis

#### Comparability of nutrient data

A Spearman’s ranked correlation was used to determine whether the log_10_ ammonia and log_10_ orthophosphate concentrations data followed similar trends and could therefore be used in parallel for flow normalisation. Additionally, a Wilcoxon signed-rank test was used to determine whether there were significant pairwise differences between log_10_ ammonia and log_10_ orthophosphate concentrations.

#### Norovirus data censoring and normalisation

Following RT-qPCR, GI and GII concentrations below LOQ or LOD were censored by assigning a value of LOQ/2 to positive samples with concentrations <LOQ and a value of LOD/2 to samples with no GI or GII detected. The GI and GII concentrations in the original wastewater samples were calculated according to Walker *et al*. (2024).

The GI and GII concentrations were normalised for flow and population according to nutrient concentrations, using a method adapted from Roberts et al. (2022). The normalisation method applied in this study differed from the Roberts et al. method by including both ammonia and orthophosphate as determinants for flow. For each sampling site, anomalous ammonia and phosphate concentrations were removed using a three-sigma outlier approach. Mean ammonia and orthophosphate levels were calculated for each sampling site with anomalous data removed.

For each sample, flow was calculated separately with log_10_ ammonia and orthophosphate concentration according to Roberts *et al*. (2022), and the mean of the two values was used as the overall flow measurement for normalisation. Where data were missing for both nutrients, the sample was excluded from further analysis. The normalised NoV concentrations were presented as gene copies per 100,000 capita (copies/100k capita).

Total NoV concentration was calculated as the sum of GI and GII concentrations after censoring. Analyses were carried out using log_10_ flow and population-normalised GI and GII and total NoV data.

#### Differences in genogroup concentrations (ΔG)

Differences in GI and GII concentrations were modelled by comparing fitted smooths in factor-smooth interactions of simple Generalized Additive Models (GAMs) (Simpson, 2017). The models may be represented as:

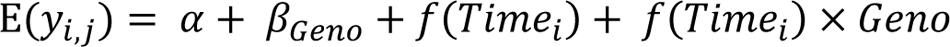

where *y*_*i,j*_ is the log_10_ concentration at time point *i* of genogroup *j*, α is the parametric effect of genogroup II ( i.e. the reference level), β_*Geno*_ is the parametric effect of genogroup I, *f*(*Time*_*i*_) is the smooth function of Time on genogroup II and *f*(*Time*_*i*_) × *Geno* is the smooth function representing the trend in difference between log_10_ GI and GII concentrations. From these models the trends in log_10_ difference between GI and GII concentrations (ΔG) were obtained using the difference smooth term *f*(*Time*_*i*_) × *Geno* offset by the β_*Geno*_ coefficient. Thin-plate regression splines were used to parametrise the smooth functions, *f*() (Wood, 2003) and models were fit in R using the mgcv package, version 1.8-41 (Wood, 2017). Plotting of modelled trends in ΔG was implemented with the gratia package, version 0.8.2 (Simpson, 2024).

#### Temporal trends in the national and regional datasets

To analyse temporal trends in NoV concentrations and ΔG in wastewater across England, data were aggregated by two-weekly periods starting on 27^th^ May 2021. All periods with fewer than 100 data points in the national dataset were removed from further analysis to ensure robustness of the data analyses. The periods were further split by region, and all regional aggregates with fewer than 8 data points were removed from further analysis. The log_10_ mean averages were calculated for GI, GII, total NoV and ΔG from all sites for each of the national and regional two-weekly aggregates. Upward and downward trends in NoV levels were calculated using a relative strength index (RSI) with the TTR package (Ulrich, 2021) for R (R Core Team, 2023) according to Chan et al. (2023), using a rolling 6-week look-back period. Each RSI value was assigned a descriptive trend category according to Table 2.

**Table 2:**
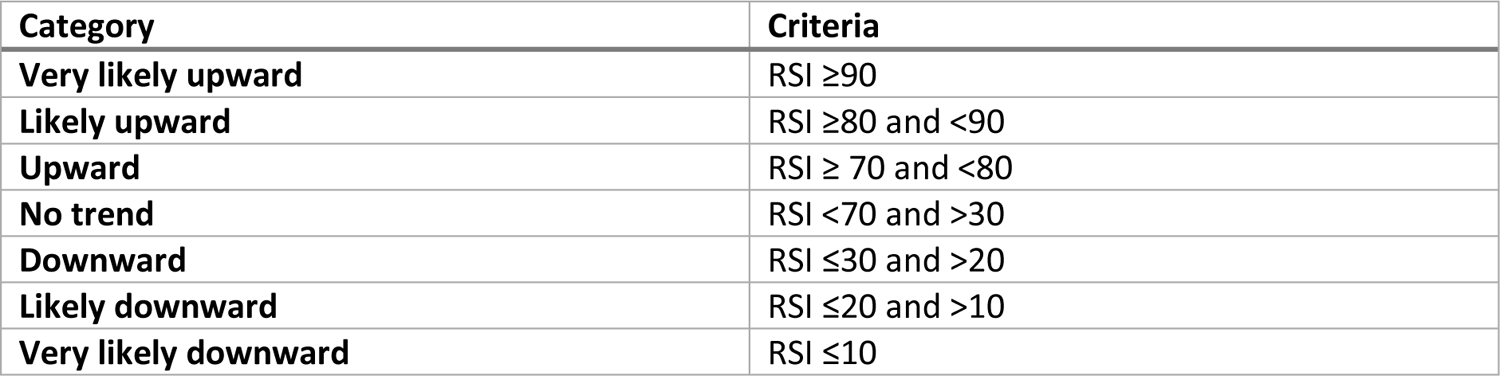
Descriptive categories for trend analysis based on a relative strength index for a rolling 6-week look back period for norovirus concentrations in wastewater.

#### Site to site analysis of norovirus data

Due to variability in the timing of sample collection dates, results were aggregated by calendar month for site specific spatiotemporal analysis. The mean log_10_ GI and GII concentrations were calculated per site and per month for those sites with multiple sample dates within the same month. Sites that did not have at least one sample result per genogroup per month were removed from the analysis. This left 140 sites with sample results for NoV GI and GII from June 2021 to March 2022, yielding a total of 2,800 sample results.

The minimum distance to ensure each site would have at least one neighbour (and, therefore, be included in the local indicators of spatial association (LISA) assessment), was determined, and site locations were evaluated for spatial autocorrelation at that distance using a modified Ripley’s K analysis (Besag, 1977; Ripley, 1977). LISA assessment of the site Log_10_ results was carried out using a Local Moran’s I analysis (Anselin, 1995) to identify both clusters and outliers of high or low values. All of these analyses were carried out using tools available within ArcGIS Pro version 2.9.5 (ESRI, 2022).

#### Comparison of wastewater data against clinical case numbers

National weekly NoV case numbers were taken from the UKHSA National NoV and rotavirus bulletin reports (UKHSA, 2023b). The weekly case numbers were aggregated into two-weekly time periods starting on 24^th^ May 2021, and the sum of the cases was calculated for each period. Total NoV concentrations in wastewater were aggregated by two-weekly periods starting on 24^th^ May 2021. The distributions of the aggregated two-weekly case numbers and log_10_ average NoV concentrations were checked for normality with the Shapiro-Wilk test and then compared by linear regression in R.

## Results

The data from this study can be freely accessed from (David I. Walker *et al*., 2024).

### Ammonia and Orthophosphate Concentrations

Figure 2 shows the distribution and comparisons of the concentrations of the nutrients ammonia and orthophosphate in wastewater samples. Of the 3,232 samples, 3,220 had associated ammonia or orthophosphate data and were used in further analysis. Three additional samples were removed from further analysis following 3σ rule outlier analysis for ammonia and orthophosphate. According to Shapiro-Wilk tests, the nutrient concentrations were not normally distributed (p<0.001 in both cases). A non-parametric Wilcoxon signed-rank test showed that the pairwise ammonia and orthophosphate concentrations for samples were significantly different (p<0.001), with ammonia concentrations exceeding the orthophosphate by a mean average of 1 log_10_ mg/l with a standard deviation of 0.16 log_10_ mg/l. A Spearman’s rank correlation test between log_10_ ammonia and log_10_ orthophosphate concentrations showed a significant correlation with ρ = 0.811 and p<0.001.

**Figure 2:**
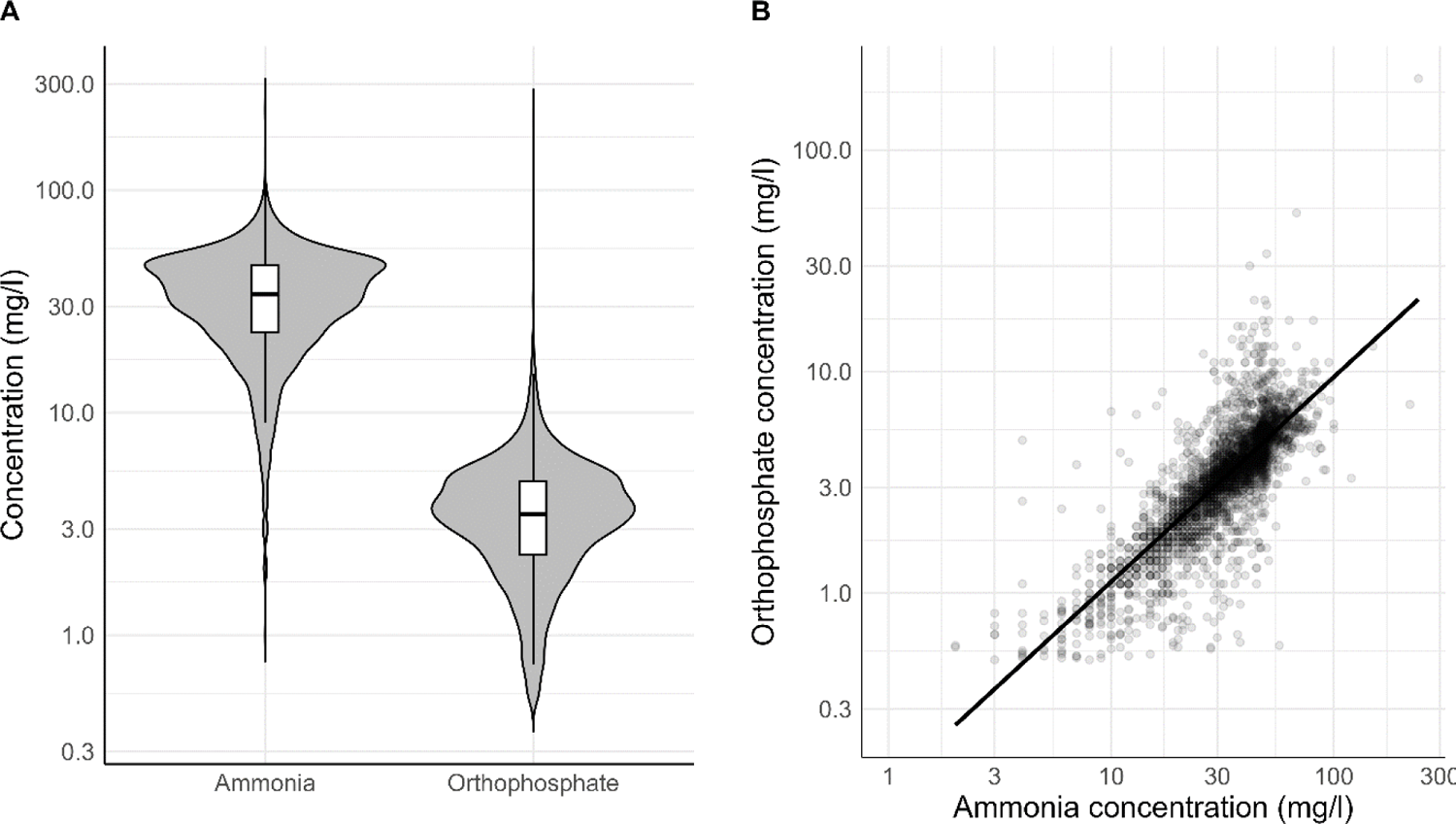
(A) Violin and box plots on a log10 scale, showing the distributions of the ammonia and orthophosphate data for 3,220 wastewater samples collected across England. Grey shaded areas are frequency of data. White boxes are the interquartile range, horizontal lines represent the median. (B) Scatter plot showing ammonia concentrations vs orthophosphate concentrations on a log10 scale. Black line represents a line of best fit on a log10-log10 scale.

### Norovirus concentrations

The LOD_95_ and LOQ for the duplex RT-qPCR assay were calculated as 9.5 gc/reaction and 11.4 gc/reaction respectively for GI and 6.7 and 16.3 gc/reaction respectively for GII. Table 3 shows the number of NoV positive samples and number of samples that were either non-detects or <LOQ for each genogroup.

**Table 3:**
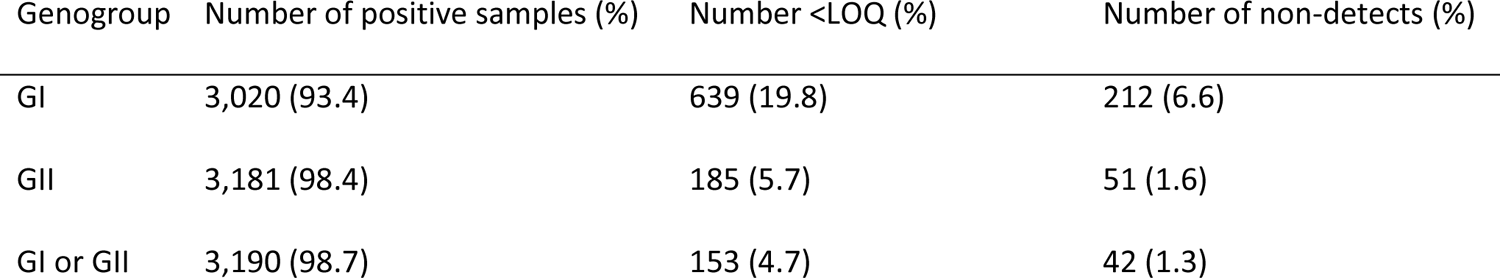
A summary of the number of samples that had no norovirus genogroup I and II detected or were below the limit of quantification (LOQ). This is expressed, in brackets, as a percentage of the 3,232 samples.

Figure 3 shows the distributions of the GI and GII concentrations following censoring and normalisation. Shapiro-Wilk tests indicated that the GI and GII data were not normally distributed. A non-parametric Wilcoxon signed-rank test indicated that there were significant differences in pairwise GI and GII concentrations across the national dataset (P<0.001). The median log_10_ GI and GII gene copies per 100k capita were 4.27 and 4.96, respectively.

**Figure 3:**
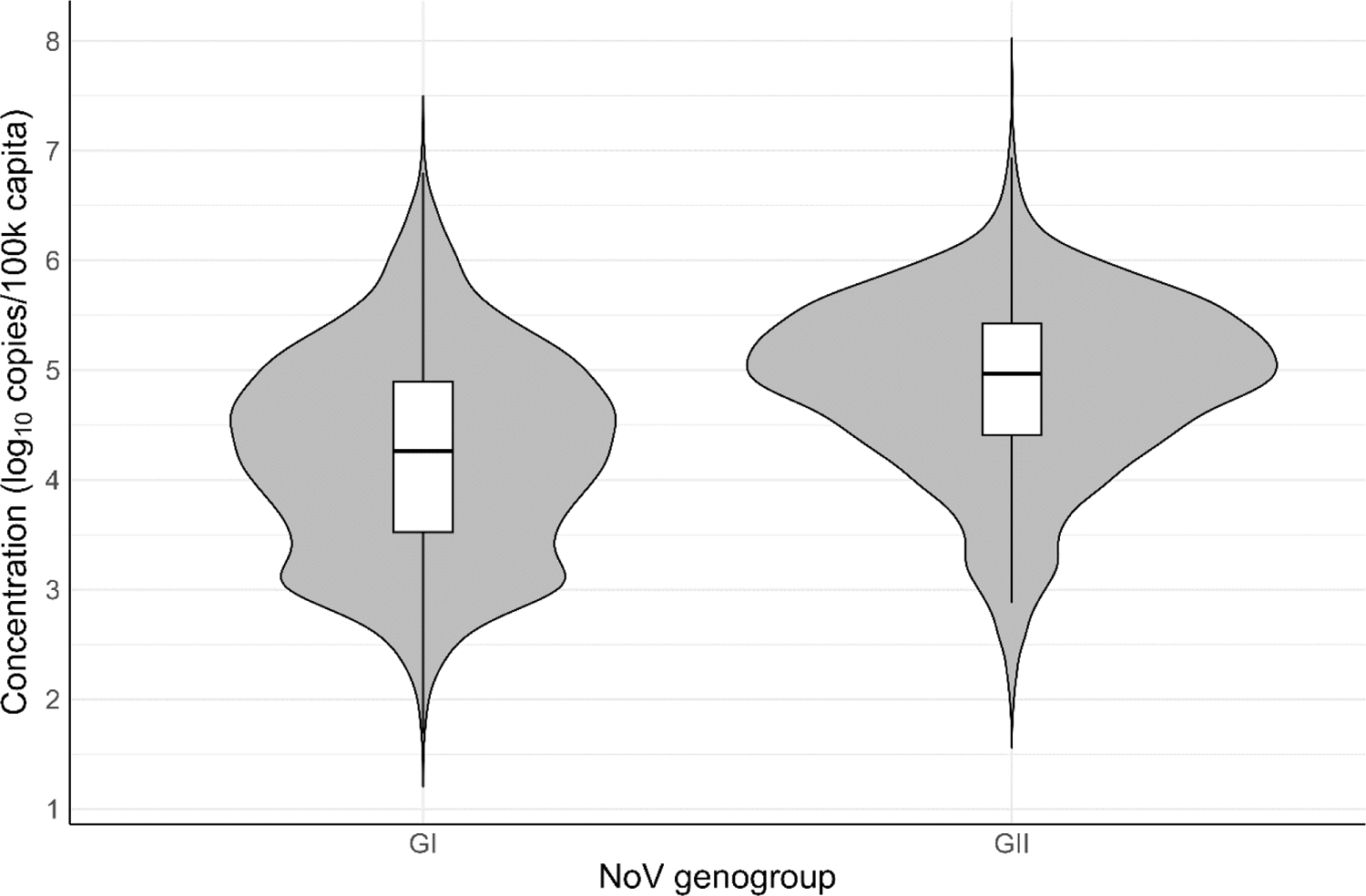
Violin and box plots showing the distributions of the log10 concentrations of norovirus genogroups I (GI) and II (GII) for 3,220 wastewater samples collected across England. The grey shaded areas represent the frequency of data. The white boxes represent the interquartile range, and the horizontal lines represent the median.

### Temporal Tends in NoV Concentrations in wastewater

Figure 4 shows the overall trend in NoV concentrations in wastewater samples across England throughout the sampling period (27^th^ May 2021 and 30^th^ March 2022) as two-weekly mean concentrations for GI, GII and total NoV with the RSI trends for those periods. During the summer of 2021, there was an upward trend in both GI, GII and total NoV levels within wastewater across England. This appeared to coincide with the staggered removal of NPIs (lockdown measures) for COVID-19, which mostly ended on 19^th^ July 2021 (UK Government, 2021b). This was followed by a period up to late December 2021 where the GII and total NoV levels showed little overall change. However, there was a downward trend in GI in September 2021. In January 2022, there was a downward trend in GII and total NoV, which appeared to coincide with the introduction of further NPIs for COVID-19, referred to as “Plan B” (UK Government, 2021a). This was followed by a marked upward trend in GI levels in February and March 2022, which was reflected by the upward trend in total NoV. During this period, the GII levels also trended upward.

**Figure 4:**
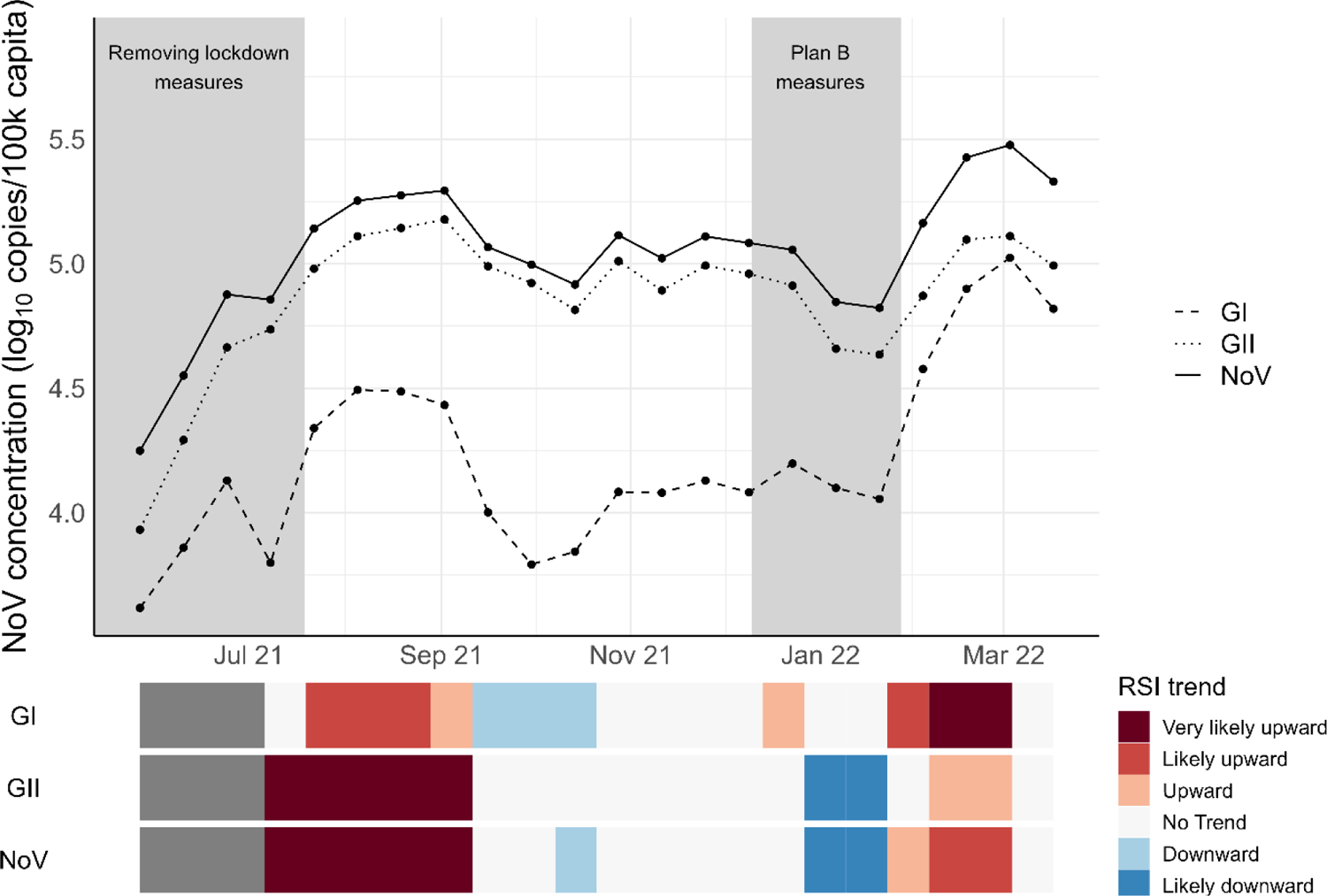
The log10 national two-weekly national average concentrations of norovirus genogroups I and II, and total norovirus (GI + GII) from May 2021 to March 2022. The grey shaded areas show periods of non-pharmaceutical interventions for controlling outbreaks of COVID-19 in England. The relative strength index (RSI) is also shown for each measure, indicating the strength of the change in concentrations over a 6-week rolling look-back period.

### Temporal Trends in Differences Between Genogroup Concentrations (ΔG)

Figure 5 shows the national average log_10_ differences in GI and GII concentrations measured in the wastewater samples (ΔG) in two-weekly sampling periods. In this case, a value below zero indicates a higher concentration of GII relative to GI. While ΔG appeared to decrease in the first half of the study, ΔG increased over the later period (from October 2021). This indicates an increase in the proportion of GI relative to GII. Figure 6 shows the regional average log_10_ differences in GI and GII concentrations measured in the wastewater samples (ΔG) in two-weekly sampling periods. A trend towards higher GI accounting for a higher proportion of the total NoV was particularly apparent in the north of England towards the end of the sampling period. All three northern regions (Yorkshire and Humber, North West and North East) had a ΔG exceeding 0 for several weeks in 2022.

**Figure 5:**
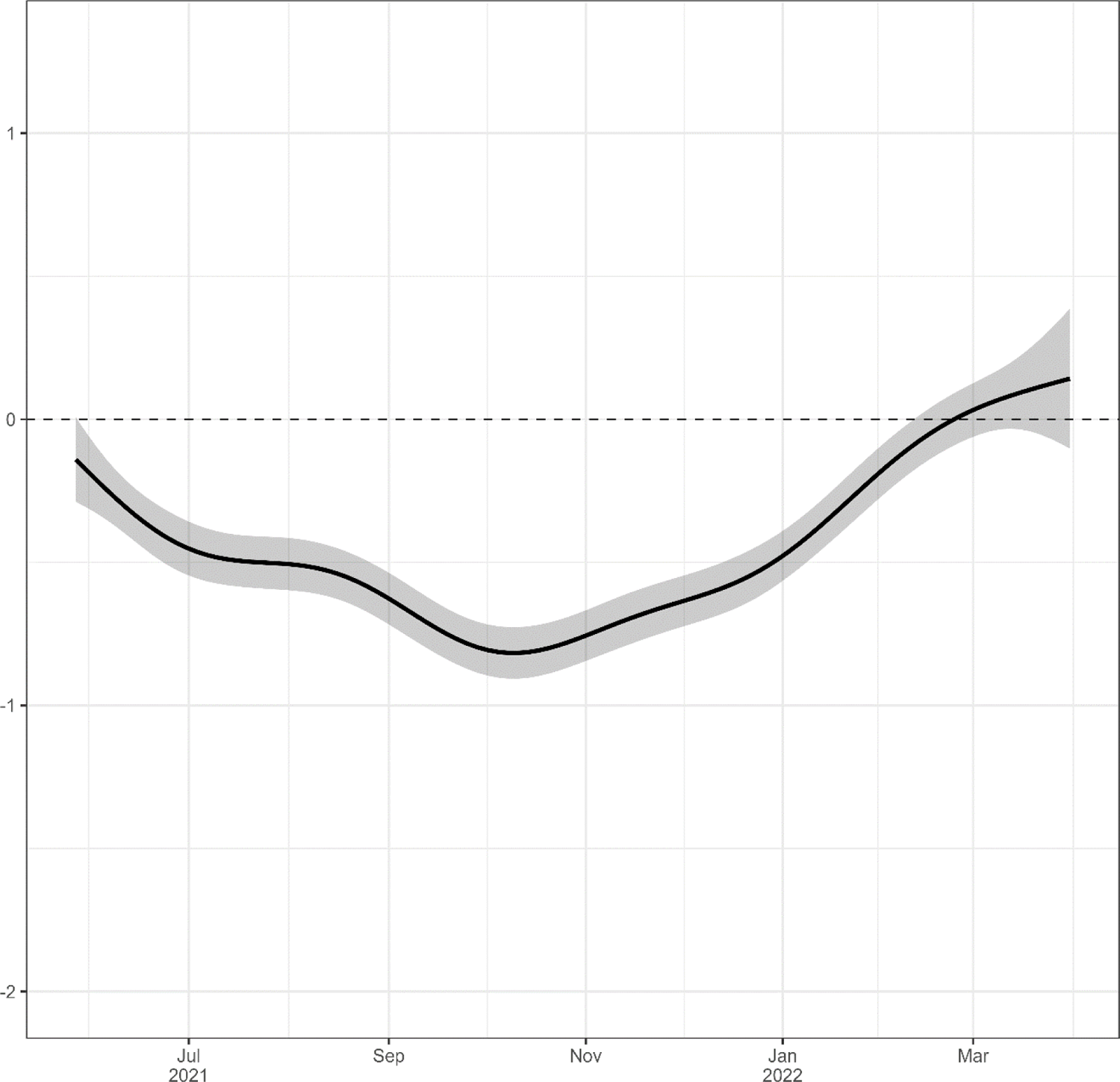
Two-weekly average log10 differences between norovirus genogroups I and II (ΔG) across England from May 2021 to March 2022. The grey area represents approximate, 95% pointwise confidence intervals.

**Figure 6:**
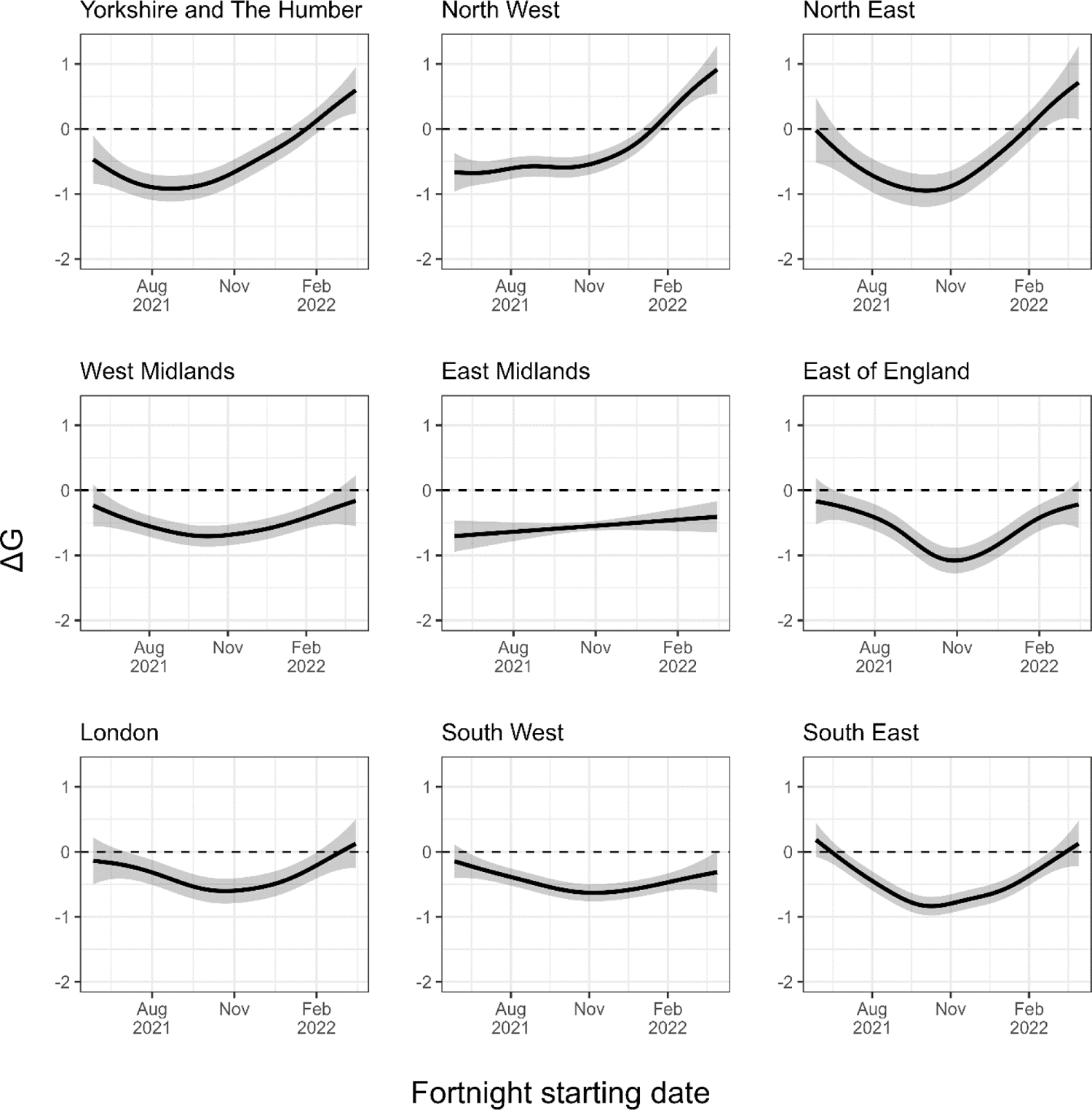
Two-weekly average log10 differences between norovirus genogroups I and II (ΔG) across the nine regions of England from May 2021 to March 2022. The grey area represents approximate, 95% pointwise confidence intervals.

### Site to Site Analysis of Norovirus Data

The minimum distance to ensure each site would have at least one neighbour was approximately 90 km. The modified Ripley’s K analysis found no significant spatial autocorrelation at this distance.

Monthly results of the Local Moran’s I analysis for June 2021, September 2021, December 2021 and March 2022 are shown in Figure 7 for GI and Figure 8 for GII. Significant clusters and outliers were identified for both GI and GII, but these varied between genogroups and over time. High-high clusters were seen for GI in the south of England in June 2021. In September 2021, high-high clusters of GI were prevalent in the South West, and were no longer seen in the South East. During this time, there were several GI high-low outliers in the north of England as well as several low-low GI clusters. In December 2021, there were high-high clusters of GI in the North West. By March 2022, there were high-high clusters of GI across the north of England. For GII, there were low-low clusters in and around London in June 2021, with some high-low outliers. Low-low GII clusters and high-low GII outliers persisted in and around London and the South East in June 2021, and by March 2022, these were no longer present. In June 2021 and March 2022, there were low-low GI clusters and high-low GII outliers in the north of England. High-high GII clustering was seen at several sites in the South West in September 2021, but were no longer present by March 2022.

**Figure 7:**
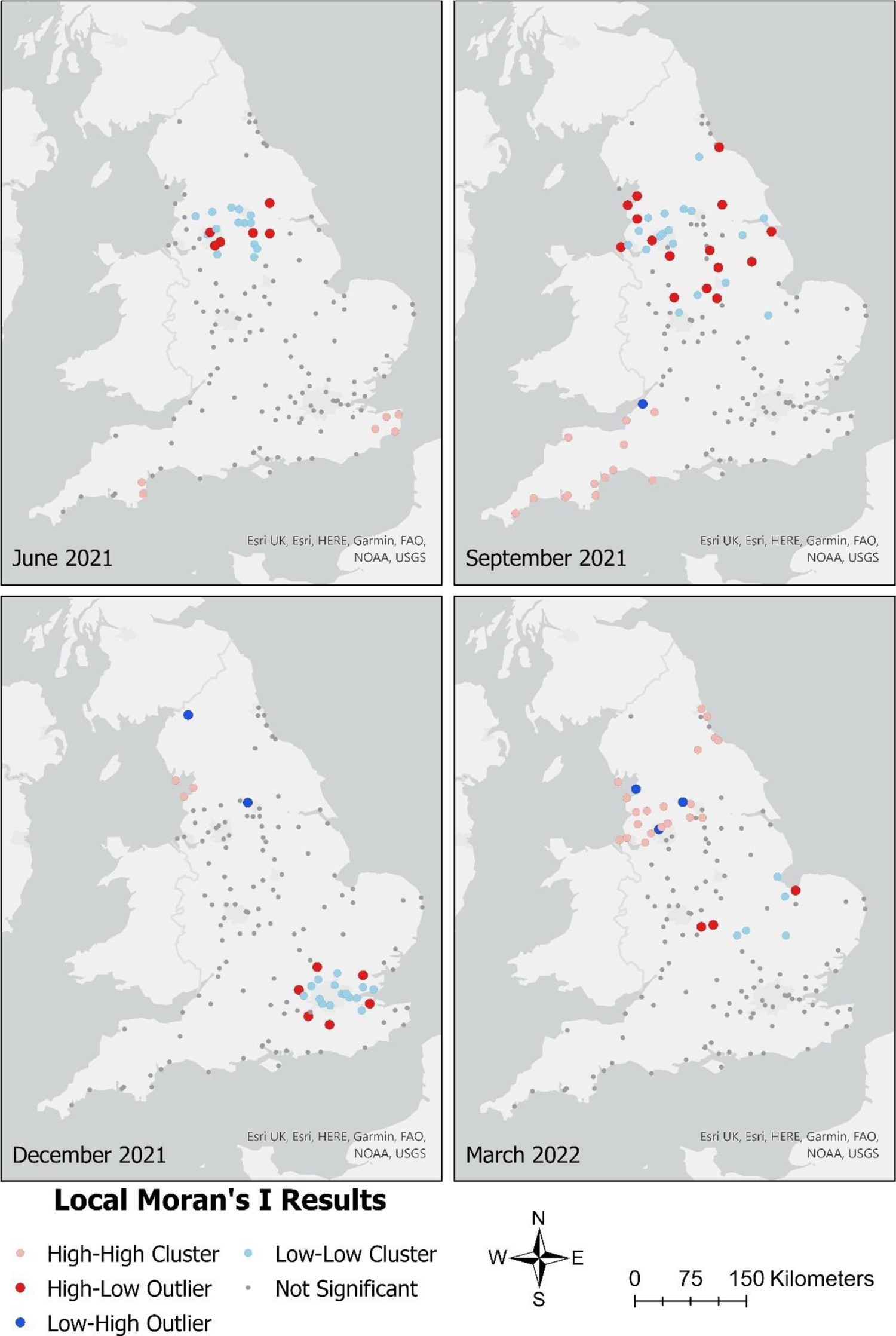
The location of the sampled sewage treatment works and the results of Local Moran’s I analysis on log10 concentrations of norovirus genogroup I from these locations in June 2021, September 2021, December 2021, and March 2022

**Figure 8:**
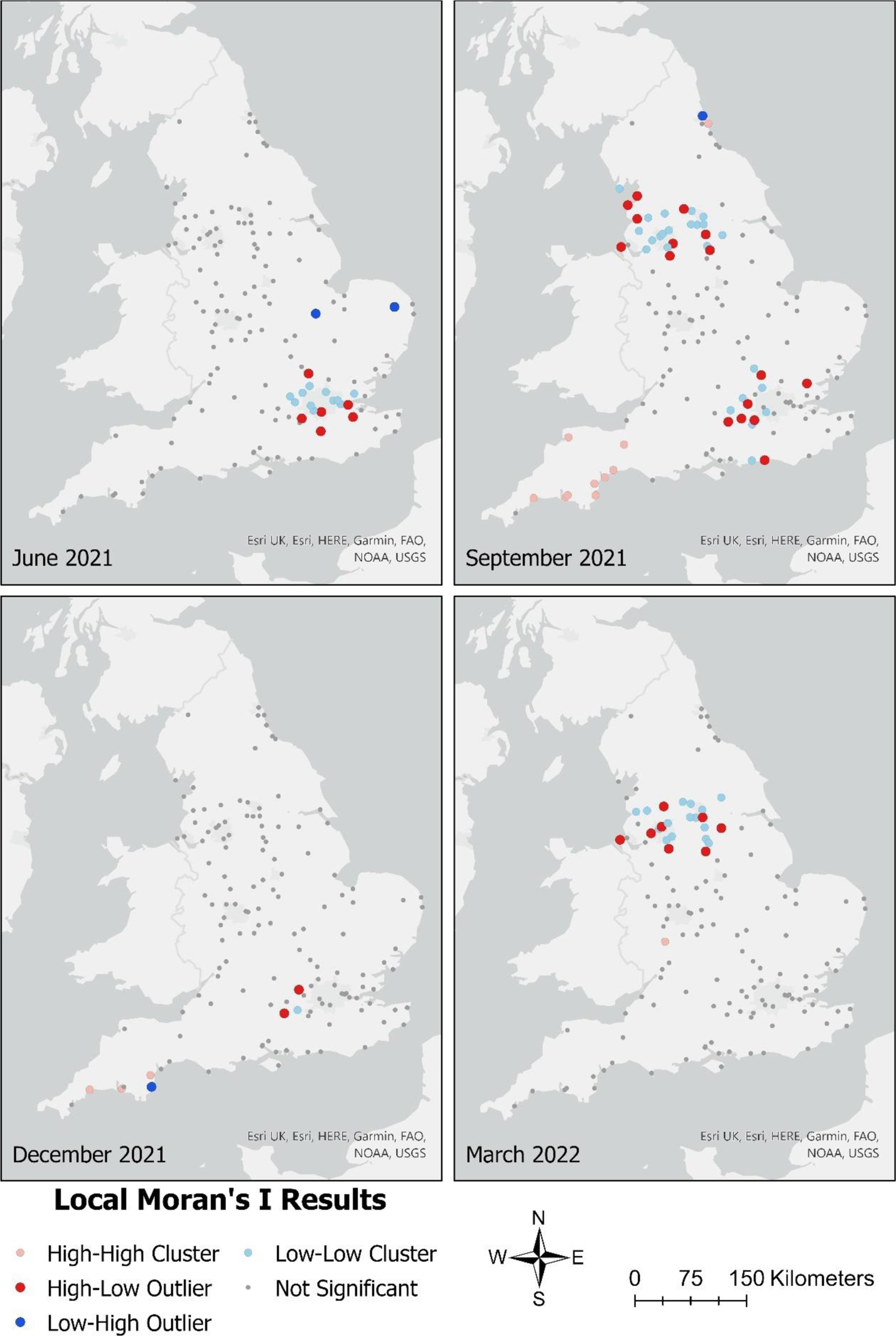
The location of the sampled sewage treatment works and the results of Local Moran’s I analysis on log10 concentrations of norovirus genogroup II from these locations in June 2021, September 2021, December 2021, and March 2022

### Comparison of wastewater data against clinical case numbers

Linear regression analysis showed a significant (p<0.001) linear relationship between wastewater data vs. clinical case numbers, with a R² of 0.614. This relationship was stronger when both measures were compared on a log_10_ scale (R² = 0.720, p<0.001). To determine the effect of flow normalisation on the relationship between log_10_ NoV concentration in wastewater and log_10_ reported case numbers, the linear model was repeated using NoV concentrations not adjusted for flow. The linear relationship remained significant (p<0.001) but was weaker (R²=0.634). All further results presented are for flow and population-normalised data.

Figure 9 shows the relationship between the two-weekly log_10_ NoV case numbers and the two-weekly national average log_10_ NoV concentrations in wastewater. In Figure 10, the NoV concentrations in wastewater are compared with case numbers over the sampling period, showing a general consensus between concentrations in wastewater and case numbers over time, but also indicating times where that relationship was weaker.

**Figure 9:**
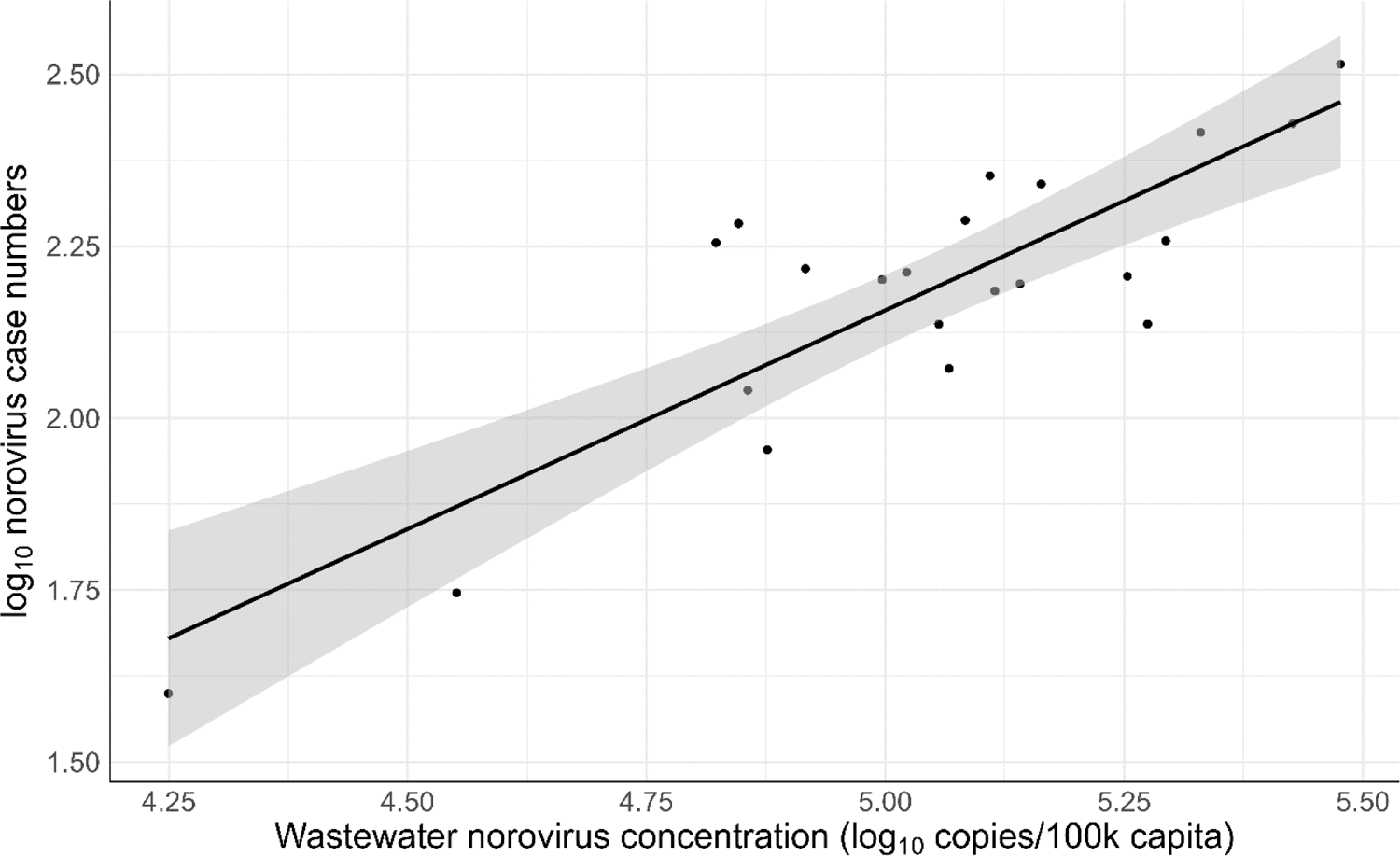
Comparison of log10 national average norovirus concentrations in wastewater against the log10 number of confirmed norovirus case numbers as reported by UKHSA. Black line represents the linear model, with the grey shaded area representing 95% confidence intervals.

**Figure 10:**
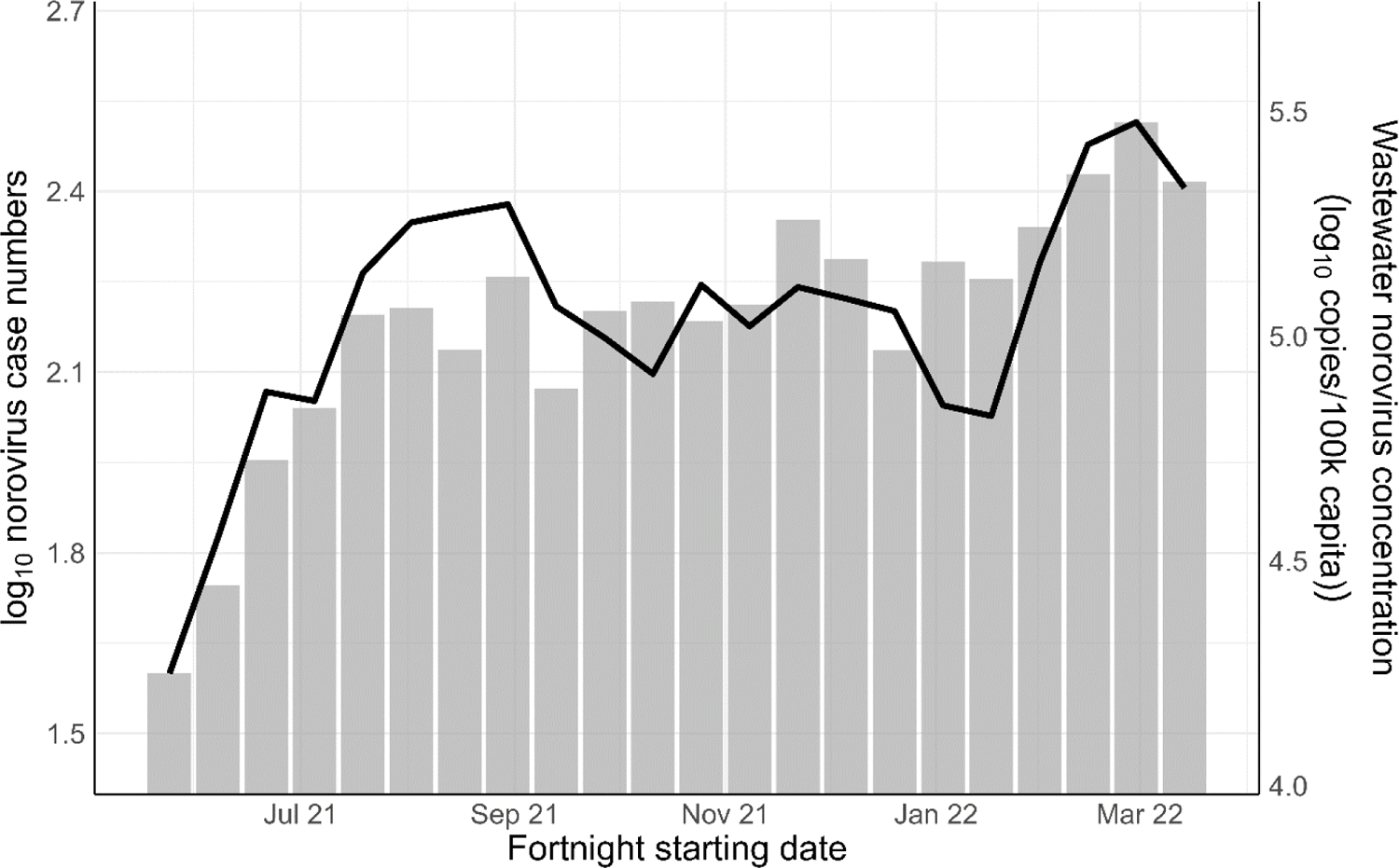
Time series comparison of log10 national average norovirus concentrations in wastewater and the log10 number of confirmed norovirus case numbers as reported by UKHSA. The black line is the concentrations in wastewater and the grey bars are the number of cases. Note the different scales of the two y-axes.

## Discussion

In this study, the trends in NoV concentrations in wastewater across England were assessed. As in other studies on viruses in wastewater (Morvan *et al*., 2022; Scorza *et al*., 2022), an estimate of flow and population size was applied as a normalisation factor on the NoV concentration data. For these samples, ammonia and orthophosphate data were previously collected as part of the Environmental Monitoring for Health Protection (EMHP) programme for wastewater surveillance of COVID-19 in England (Sweetapple *et al*., 2023). These measures were found to correlate significantly, which supports their use in parallel with each other. It should be noted, however, that potential infiltration of land runoff and other sources of non-human faecal material into the sewerage network, which may itself be influenced by variations in rainfall and other environmental factors, may also cause variability in nutrients (Murphy *et al*., 2015; Beheshti and Sægrov, 2018). Such factors could lead to misrepresentation of flow, thereby leading to errors in normalisation of the norovirus concentration data. Other studies have used different factors to normalise viral loads in wastewater, including concentrations of faecal indicator organisms (Zhan *et al*., 2022; Langeveld *et al*., 2023), electrical conductivity (Langeveld *et al*., 2023), and other chemicals (Hsu *et al*., 2022). However, there is currently no consensus on the most reliable measure, and it is likely that the best indicator will vary across locations with different geographies and different sewerage infrastructures. Nevertheless, reanalysis of virus data from this study without the nutrient-based adjustment for estimated flow, showed a diminished correlation between wastewater NoV concentration and case numbers, as reflected by a reduction in the strength of their linear relationship. This indicates that while the nutrient-based normalisation may not be perfect, such adjustments may help to improve the insights gained from virus concentrations in wastewater.

The very high prevalence of NoV in wastewater (99% of samples) was not unexpected given the high prevalence of norovirus in communities (Tam *et al*., 2012). According to UKHSA reports from the same period (UKHSA, 2023b) GII accounted for 90% of the reported clinical cases in 2020 and 2021, so this approximately 1 log_10_ difference between GI and GII might be expected to be reflected to in wastewater, assuming similar faecal shedding rates for each genogroup. This study did not find this degree of overall difference between GI and GII concentrations, the average GII concentrations being approximately 0.7 log_10_ greater than GI, and this figure varied considerably across the sampling period.

Similarly, studies of NoV prevalence in bivalve molluscan shellfish (BMS) have shown GII to be present at higher concentrations in most cases (Lowther *et al*., 2018; EFSA, 2019). However, this difference found in BMS may be confounded by differences in binding kinetics of NoV genogroups and genotypes in BMS digestive tracts (Maalouf *et al*., 2011; Morozov *et al*., 2018) A systematic review and meta-analysis of 26 studies by Huang et al. (2022) found that NoV concentrations in wastewater were typically higher in spring and winter than in summer and autumn. A similar trend has been shown in both clinical case numbers (UKHSA, 2023b) and BMS (Lowther *et al*., 2018; EFSA, 2019). Unlike these studies, the data presented here do not show these normal seasonal trends. The sampling timeframe for this study encompassed two periods of highly unusual national restrictions in England due to the Covid-19 pandemic. As such the data and trends found here were likely not reflective of a more typical period without these restrictions. Despite this, analysis of the trends under these unusual circumstances revealed some patterns of potential interest. For example, an upward trend in NoV concentrations during the first half of 2021 suggests increased person-to-person transmission as national restrictions were lifted during this period.

During late autumn and early winter of 2021, when NoV levels would usually be expected to increase, no such increase was found and in fact, a likely downward trend in GII and overall NoV concentrations was observed during the winter period. That may have been a result of further national restrictions, known as “Plan B”, that were put in place to limit the spread of the Omicron variant of SARS-CoV-2. Notably, levels of NoV in wastewater increased again as these restrictions were lifted. This may have led to a delay in the usual winter peak in NoV that was otherwise limited.

Overall, GII remained higher than GI over the sampling period as reflected by the higher average concentration of GII as previously discussed. However, investigation of the log_10_ differences in GI and GII over time revealed that this relationship did not remain consistent. The first half of the sampling period was characterised by a general decrease in the concentrations of GI relative to GII. But from October 2021 onwards, GI concentrations consistently increased relative to GII at a national level.

Further investigation of this pattern at a regional level showed that this was markedly different across the country. In the northern regions of Yorkshire and The Humber, North West and North East, GI became the dominant genogroup by the end of the study period, suggesting a localised spread of GI at that time. This was in contrast to South West, East Midlands and West Midlands regions, where no strong changing trends in GI and GII levels were evident.

Local Moran’s I analyses indicated potential localised outbreaks of both GI and GII across England. The high-high GI clusters seen in the north of England in December 2021 and March 2022 may show that there was an outbreak of GI in the North West, in the Morecambe Bay area. By March 2022 this had spread eastward to cover a large part of the north of England. Also during March 2022, there were several low-low clusters of GII in the north of England that may indicate that GI largely replaced GII as the dominant genogroup of norovirus in human populations in these regions at that time. The large number of high-low outliers seen for both GI and GII may be an indication of localised outbreaks that were contained due to the nature of the communities or the potential start of a wider outbreak. This shows that Local Moran’s I analysis, when used in wastewater surveillance, can be useful in investigating the spread of pathogens through communities. It should be noted however, that in the context of the current study, sampling sites were selected with a bias towards larger population sizes, and primarily in densely populated areas. It therefore cannot be assumed that the observed trends hold true for the whole population.

To determine whether NoV concentrations in wastewater could predict a disease burden from NoV, the national average log_10_ total (GI+GII) concentrations in wastewater were compared with national total log_10_ reported case numbers for two-weekly periods throughout the survey period. The significant linear relationship between these measures indicates that concentrations of NoV in wastewater are at least somewhat representative of the disease burden. However, it should be noted that neither wastewater surveillance nor clinical case numbers are likely to give a completely accurate picture of the true number of national cases of NoV for various reasons, including sampling strategy, environmental factors and reporting of cases. The second Infectious Intestinal Diseases Study (IID2) indicated that NoV cases are currently underestimated at a rate of approximately 300 community incidences per one reported incidence (Tam *et al*., 2012). Additionally, Ondrikova et al. (2021) found substantial variability in NoV reporting between English regions and age groups, and Douglas et al. (2021) found that NoV reporting rates were significantly impacted by national lockdown measures. These factors are likely to result in significant bias in data sets when attempting to analyse trends.

Wastewater-based surveillance of pathogens attempts to minimise this bias by introducing a non-invasive system and reducing the likelihood of demographic variation in estimates of disease burden. However, the use of wastewater-based surveillance introduces its own biases. Such biases were discussed in detail in the context of SARS-CoV-2 by Wade et al. (2022), and include factors such as population characteristics, sewerage network characteristics, sampling strategy and sample analysis. While biases arising from sample analysis methods may be at least partially resolved by optimising bench methods, the other factors will be more difficult or impossible to resolve. Population statistics, and environmental and infrastructure data should, therefore, be considered for their value in supporting, validating, and optimising surveillance programmes that use wastewater-based data sources. Using variations in sewerage network characteristics as an example, it is common to normalise pathogen load by flow rate and population size (Hsu *et al*., 2022; Zhan *et al*., 2022; Langeveld *et al*., 2023), as applied in this study, where relative flow was estimated using deviations from average nutrient concentrations on a sample-by-sample basis. As discussed previously, the inclusion of this type of normalisation strengthened the relationship between clinical case numbers and NoV concentrations in wastewater. This indicates however, that discrepancies between reported case numbers and viral concentrations in wastewater may, in part, be a result of imperfect normalisation methods. Due to the likelihood of biases in both reported case data and wastewater-based data, it is important to recognise that data derived from wastewater are likely to have the greatest value as part of a multi-faceted approach to pathogen surveillance, rather than as the sole data source. Despite the different biases inherent in both data sources, it is reassuring that there was a strong correlation between norovirus levels in wastewater and reported clinical cases.

The results of this study show the value of wastewater monitoring as a non-invasive tool for surveillance of enteric diseases such as NoV in human populations. This will be particularly valuable where reporting of cases is low relative to the estimated number of cases in the community, and where other factors that may bias reporting are dominant.

Further optimisation of the sampling programme is required for long-term surveillance systems. This must include consideration of the most cost-effective number and location of surveillance sites within any programme, further development of tools for normalisation of viral loadings, improved characterisation and optimisation of sample analysis methods, and continued work to develop models for the integration of wastewater data within existing clinical datasets.

## Conclusions

- The unusual circumstances due to COVID-19 related national restrictions appear to have reduced the levels of norovirus circulating in communities and therefore wastewater.
- Variations in the national, regional, and local levels of norovirus genogroups I and II indicated the potential for use of wastewater-based methods for detecting outbreaks.
- The high level of conformity between norovirus levels in wastewater and reported clinical cases suggests that wastewater-based monitoring could provide a valuable source of information for understanding the epidemiology of norovirus in England.
- Continued optimisation of sampling programs, considering cost-effective surveillance sites, improved normalization methods, and integration with clinical datasets, is necessary for long-term effectiveness.

## Data Availability

All data produced are available online at https://doi.org/10.14466/CefasDataHub.148

## Acknowledgements

We would like to thank all the members of the EMHP programme for the collection and processing of all of the samples during the COVID-19 pandemic. We would like to thank Dr Irene Bossano for her work to provide the sub-set of samples required for this study. We would also like to thank Dr Sarah Alewijnse for maintaining the data holdings for this project.

